# Applications of Synthetic Data Integration for Deep Learning for Volumetric Analysis and Segmentation in Thoracic CT Imaging

**DOI:** 10.1101/2024.10.30.24316446

**Authors:** Ali Zeyrek, Sergio M Navarro

## Abstract

This study presents a framework for processing Digital Imaging and Communications in Medicine (DICOM) medical imaging data by integrating synthetic objects for volumetric analysis and simulation for applications in assessment of computed tomography (CT) imaging used in thoracic surgery. Functions are designed to generate synthetic objects including geometric shapes such as spheres, cubes, rectangular prisms, cylinders, and blobs with known volumes. Validation is performed through test functions to ensure accuracy and consistency. Additionally, the use of UNet models for segmenting various chest pathologies, such as hemothorax and pneumothorax, as well as organs, is demonstrated. The created framework is used to generate synthetic data to address the scarcity of publicly available hemothorax CT imaging data. Models achieved high performance, assessed by various metrics. The framework and models provide a robust tool for data augmentation and analysis in medical imaging, potentially enhancing clinical decision-making and supporting research in thoracic surgery and related fields.

## 1. Introduction

Understanding the volume of hemothorax and other various lesions, and measuring their volumes is crucial for determining a diagnosis. This paper details a general purpose framework for synthetic data generation that processes DICOM medical imaging data, creates synthetic objects such as spheres and blobs, and estimates their volumes using the Monte Carlo method. Using this framework, this paper illustrates and explains the training of segmentation models for chest organs, hemothorax and pneumothorax.

## 2. Methods

### 2.1 Reporting Guidelines

The study followed appropriate research reporting guidelines, including relevant EQUATOR research reporting checklists and other pertinent reporting as applicable. Data sources are referenced and only publicly available data sources were used for the study, with Institutional Research Board exemption granted based on checklist review. This work used ONLY simulated data or ONLY data that was openly available to the public before the initiation of the study.

### 2.2 Synthetic Hemothorax Data Framework

Synthetic hemothorax data framework enables us to overcome the scarcity of publicly available hemothorax data. Using the tool we created, we can synthetically place specific 3D objects in DICOM series, on any arbitrary coordinates and dimensions, with flexible options for setting object boundaries and Hounsfield values.

### 2.3 Data Loading

Patient data is loaded and organized using the `load_patient_data` function, which retrieves DICOM series from a specified folder. Function preserves metadata and reads multiple series if there are for a patient.

### 2.4 Object Declaration

In the class `Object`, there are functions for multiple objects: sphere, quadrilateral, cylinder, blob. Besides the required arguments for all, such as the position (z,y,x), HU baseline value, HU noise amplitude and object noise, shapes may also require distinct arguments. The purpose of the functions are essentially to check whether a certain point is within the bounds of the object.

#### 2.4.1 Sphere

The `Sphere` function creates a sphere function based on the given radius and center coordinates. Function checks whether the distance *r* from a point (*x, y, z*) to the center (*x*_0_, *y*_0_, *z*_0_) is within a given radius *R*, with added random noise from ‘obj_noise‘. This effectively defines a noisy spherical boundary around the center point.

#### 2.4.2 Prism

In addition to regular arguments, the `Prism` function requires height, length and width. Function checks whether the coordinates (*x, y, z*) fall within the 3D rectangular region centered at (*x*_0_, *y*_0_, *z*_0_), with dimensions defined by ‘width‘, ‘length‘, and ‘height‘. The ‘obj_noise‘ term introduces random variations to each boundary, also simulating noise or irregularity if requested.

#### 2.4.3 Cylinder

The `Cylinder` function requires height and radius besides regular inputs. Function checks if a point (*x, y, z*) lies within a noisy cylindrical region centered at (*x*_0_, *y*_0_, *z*_0_), with a specified radius *R* and height *h*. It considers both the radial distance from the center in the *xy*-plane and the vertical position in the *z*-axis, adding specified noise to simulate irregular boundaries.

#### 2.4.4 Blob

`Blob` function evaluates whether a point (*x, y, z*) falls within a 3D Gaussian distribution centered at (*x*_0_, *y*_0_, *z*_0_), characterized by amplitudes *A*, standard deviations (*σ*_*z*_, *σ*_*y*_, *σ*_*x*_), and a threshold. It introduces noise to the threshold, creating a “blobby” region with randomized intensity variations.

### 2.5 DICOM Utilities

For making the tool capable, there are various utilities besides object types. User can define a HU value for the object, and optionally make it noisy by giving a variability amplitude for a normal function. Also, user can define object threshold noise, which makes the bounds of the object noisier.

#### 2.5.1 Hounsfield Unit (HU)

##### Definition

In computed tomography (CT), the **Hounsfield Unit (HU)** is used as a standardized measure of tissue density. It is derived from the linear attenuation coefficients of tissues, normalized such that the density of distilled water at standard pressure and temperature is set to **0 HU**, while the air density is set to **-1000 HU**. This scale enables the differentiation of various body tissues based on their density.

The HU value is calculated from the linear attenuation coefficient (*μ*) using the formula:

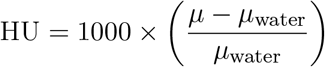

Where: - *μ* = attenuation coefficient of the tissue, - *μ*_water_ = attenuation coefficient of water.

Thus, HU values quantitatively represent tissue densities in CT images, ranging from **-1000 HU** (air) to more than **1000 HU** (bone), with other tissues like fat, muscle, and blood having values between these extremes.

##### Applying HU to Simulated Objects

When a simulated object is introduced into a CT volume, its density needs to be represented accurately in terms of Hounsfield Units to maintain the realism of the imaging. In this case, the simulated object’s HU value is defined by a user-specified mean value, typically corresponding to the expected density of the simulated tissue (e.g., **soft tissue = 30 HU, bone = 700 HU**). To introduce realistic variation, Gaussian noise is added:

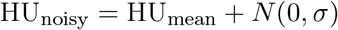

Where: - HU_noisy_ = modified HU value with noise, - HU_mean_ = mean HU value set for the object, - *N* (0, *σ*) = Gaussian noise with mean 0 and standard deviation *σ*.

The addition of Gaussian noise models the natural variability seen in real tissue densities, ensuring that the object integrates more naturally into the CT image.

##### Conversion of HU to Pixel Values

CT images typically store pixel values that are not directly in HU but require conversion using linear scaling parameters provided in DICOM metadata: **Rescale Slope** and **Rescale Intercept**. The conversion from HU to DICOM pixel values is described by:

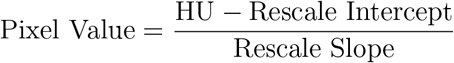

Where: - **Rescale Slope** and **Rescale Intercept** are constants provided in the DICOM header.

This conversion ensures that the modified object’s density is accurately represented in the format used by CT imaging, making it possible to simulate the appearance of the object as it would be seen in a clinical scan.

### 2.6 Object Addition

First, z,y and x dimensions of the size of DICOM series is represented as an array from 0 to n. Then, each dimension is multiplied by the PixelSpacing or SliceThickness values to get actual coordinates. Using the coordinates, a meshgrid is created. Using the meshgrid, a 5D array is created for coupling voxel centers with their corner coordinates. Corner coordinates are used for ensuring the voxel is within the object bounds completely. The voxels with all corners within the object is added to the mask.

The created objects are added into the DICOM series for the patient within the thoracic space by creating a new DICOM database. In the new database, it is indicated that slices were altered.

For a processed series, before the addition of the hemothorax, the output of the chest model is obtained first. The process is explained in later sections. This is done to ensure that the object going to be added to stay within the bounds of the lung. To achieve this, hemothorax mask and lung mask for each slice is intersected. The output is then saved as the artificial hemothorax. This ensures that artificial hemothorax is placed within the lung space.

#### 2.6.1 Volume Calculation

For calculating the volume of object, a minimum bounding box around the object is defined. Then, the volume is calculated with the following methodology.

The Monte Carlo method is used to estimate the volume of irregular objects, leveraging random sampling to determine the inclusion of points within the blob. The mathematical foundation of this approach is as follows:

Let *f* (*x, y, z*) be the blob function that defines the shape of the object, where:

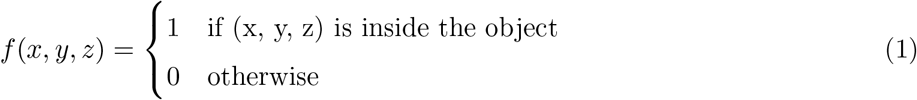

The volume *V* of the object can be expressed as a triple integral:

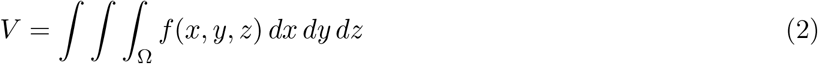

where Ω is the bounding box containing the object.

The Monte Carlo method approximates this integral by randomly sampling points within the bounding box. If we generate *N* random points (*x*_*i*_, *y*_*i*_, *z*_*i*_) uniformly distributed in Ω, the volume can be estimated as:

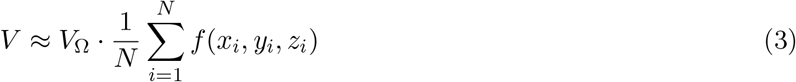

where *V*_Ω_ is the volume of the bounding box.

In our implementation, we account for the pixel spacing and slice thickness of the DICOM images. The volume of a single voxel is calculated as:

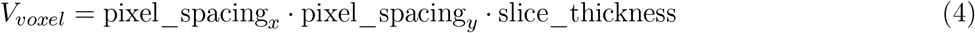

The bounding box volume is then:

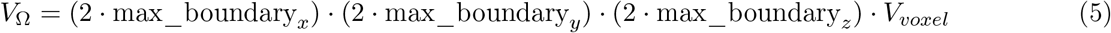

Finally, the estimated object volume is calculated as:

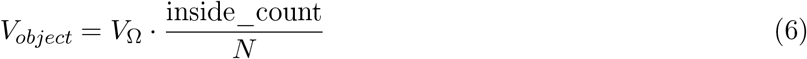

where inside_count is the number of sampled points that fall inside the object.

This Monte Carlo approach provides an efficient and flexible method for estimating the volume of irregular objects, with accuracy improving as the number of samples increases.

After the volume is calculated, it is multiplied by the ratio of pixels before and after to the lung and hemothorax mask intersection, so that volume by pixel information is accurate.

## 3. Deep Learning

Deep learning is used in multiple parts of this study.

### 3.1 Definitions and Explanations of the Components and Metrics

#### 3.1.1 Dataset

##### CT Thorax Imaging

The original dataset used for thoracic CT imaging model development is available publicly on the Harvard Dataverse (Mostafavi, 2021). The dataset consists of CT thoracic imaging of more than 1,000 patients with a confirmed COVID-19 diagnosis.

#### 3.1.2 Models

##### UNET

The UNet architecture, first proposed by Ronneberger et al., 2015, is a fully convolutional network widely recognized for its efficiency in medical image segmentation. It features a symmetric “U-shaped” design, comprising an encoder-decoder structure. The encoder progressively reduces spatial dimensions while extracting semantic features, utilizing convolutional and max-pooling layers. Conversely, the decoder employs transposed convolutions to restore spatial resolution, incorporating skip connections from the encoder to preserve high-resolution features. In this study, the UNet model was configured with the EfficientNet-B2 backbone, which is chosen for its balance between accuracy and computational efficiency. The model is optimized using pre-trained weights on the ImageNet dataset, enabling faster convergence and better performance in the segmentation of thoracic structures.

##### ResNet

ResNet (Residual Network, He et al., 2015) ( is leveraged within the UNet architecture as an encoder, with ResNet-34 as the backbone in this study. The architecture incorporates residual connections, allowing gradient flow across layers without vanishing, which aids in learning deeper representations. This characteristic is particularly beneficial for segmentation tasks, as it ensures that the features extracted from deeper layers retain relevance to the task at hand. Pre-trained weights from ImageNet were utilized for initialization, enabling the model to benefit from a broad spectrum of features learned from natural images, which in turn helps improve the segmentation of synthetic hemothorax data.

##### EfficientNet

EfficientNet (Tan & Le, 2020) represents a family of convolutional neural networks designed for better performance with fewer parameters. The scaling mechanism involves uniformly scaling depth, width, and resolution based on a compound coefficient. In this study, EfficientNet-B2 serves as the encoder within the UNet framework, primarily for its computational efficiency and robust feature extraction. EfficientNet’s depth-wise convolutions, combined with the batch normalization and Swish activation functions, contribute to the model’s ability to capture complex thoracic features while maintaining reasonable training times and memory requirements.

#### 3.1.3 Losses & Metrics

##### Dice Loss

Dice Loss (Sudre et al., 2017) is based on the Dice Coefficient, which measures the overlap between the predicted segmentation mask and the ground truth. It is defined as:

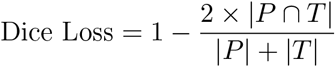

where *P* represents the set of predicted pixels and *T* denotes the set of true pixels. This formulation directly optimizes for overlap, making it suitable for tasks where accurate boundary detection is critical. By focusing on maximizing the overlap, Dice Loss aids in the precise segmentation of the regions of interest.

##### Jaccard Loss

Jaccard Loss is derived from the Jaccard Index, a metric used to quantify the similarity between two sets. It is mathematically represented as:

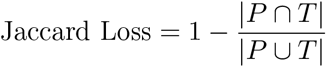

where *P* and *T* represent the predicted and true pixel sets, respectively. Jaccard Loss emphasizes the accuracy of the segmentation by evaluating the intersection relative to the union of the two sets, making it a suitable choice for assessing segmentation consistency.

##### IoU

The Intersection over Union (IoU) metric measures the overlap between predicted and true regions in segmentation tasks. It is calculated as:

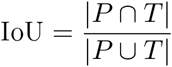

where *P* and *T* correspond to the predicted and true segmentation masks, respectively. IoU serves as a primary evaluation metric in this study, providing a clear indication of the model’s performance by quantifying the ratio of correctly predicted pixels to the total number of relevant pixels.

##### Binary Cross Entropy With Logits Loss

Binary Cross-Entropy with Logits Loss (BCEWithLogitsLoss) is a commonly used loss function for binary classification tasks, particularly effective in segmentation models where each pixel is treated as a separate binary classification problem. It combines the sigmoid activation function and binary cross-entropy loss in a single step, making it numerically stable and efficient. The formulation is:

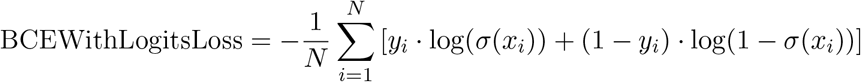

where: - *N* is the number of pixels, - *y*_*i*_ is the ground truth label for pixel *i*, - *x*_*i*_ is the predicted logit for pixel *i*, - *σ*(*x*_*i*_) represents the sigmoid function applied to the logits.

This loss function ensures accurate classification by transforming logits into probabilities in the range [0, 1] using the sigmoid function before calculating the cross-entropy. By handling raw logits directly, it avoids numerical instability, which can occur when using separate sigmoid and binary cross-entropy operations.

##### Focal Loss

Focal Loss (Lin et al., 2018) is a modification of the standard cross-entropy loss, designed to address class imbalance by down-weighting the contribution of well-classified examples and focusing more on hard-to-classify instances. It introduces a modulating factor to the cross-entropy loss that reduces the loss for correctly classified pixels, thereby prioritizing learning from harder examples. The formulation is:

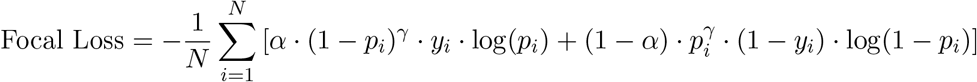

where: - *N* is the number of pixels, - *y*_*i*_ is the ground truth label for pixel *i*, - *p*_*i*_ is the predicted probability for pixel *i*, - *α* is a weighting factor to balance positive and negative classes, - *γ* is the focusing parameter, controlling the rate at which easy examples are down-weighted.

The term (1 *− p*_*i*_)^*γ*^ acts as a modulating factor, reducing the weight assigned to well-classified examples (where *p*_*i*_ is close to 1 for positive labels and close to 0 for negative labels). By doing so, Focal Loss concentrates the model’s learning on challenging pixels, making it particularly suitable for tasks with severe class imbalance, such as medical image segmentation. The parameters *α* and *γ* can be adjusted to control the balance between classes and the focus on hard examples.

### 3.2 Lung, Heart and Trachea Segmentation

Organs and important parts of thorax is segmented for bounding the thorax area. Bounding the area provides many benefits, such as estimating total volume of thoracic cavity and obtaining a boundary mask for limiting added synthetic objects.

#### 3.2.1 Data & Preprocessing

The datasets used for lung, heart, and trachea segmentation are available on Kaggle (Konya, 2021). The dataset consists of 16,708 slices of lung region with corresponding masks. Images were resized to 512×512 and normalized. 0.8 to 0.2 ratio was used for train and validation sets.

#### 3.2.2 Model & Training

A Unet with encoder of EfficientNet-B2, with weights trained on ImageNet dataset were used. Model takes data as a grayscale image, and outputs a 3 channel grayscale image. Each channel corresponds to one feature: lung, heart and trachea. Augmentations such as rotation and scaling were applied during training. Sigmoid function is applied to get probabilities from model output, and by selecting the probabilities with more than the threshold of 0.5, a binary mask is acquired. 5 fold cross validation was used.

### 3.3 Hemothorax Segmentation & Volume Estimation

#### 3.3.1 Data & Preprocessing

With the synthetic data framework, objects with random parameters were inserted in the DICOM files of the patients. Series with artificial objects were converted to .png. Out of 5250 images, 1063 had at least one artificial object added.

#### 3.3.2 Model & Training

For training, a Unet model with ResNet34 as the encoder is used. Pretrained weights from ImageNet dataset for encoder is used. For loss function, BCEWithLogits Loss and Dice Loss are used combined, with equal weights. Adam is selected as the optimizer. During training, augmentations were applied such as scaling and contrasting. 5 fold cross validation was completed. Testing was done with unseen synthetic images.

#### 3.3.3 Volume Estimation

To estimate the volume of the hemothorax, a separate linear regression model is used for a rough volume per pixel ratio. This data is obtained by running the artificial hemothorax code multiple times. Because that the actual volume of the object added is known, this provides a sufficient ground truth. This must be done only once at the beginning for each type of machine. The coefficients can be used for any scan after the process is done.

### 3.4 Pneumothorax Segmentation

#### 3.4.1 Data & Preprocessing

For pneumothorax segmentation, the SIIM-ACR Pneumothorax Segmentation Competition data was used (Zawacki et al., 2019). The dataset has 10,675 X-Ray scans after preprocessing, annotated by medical experts from Society for Imaging Informatics in Medicine (SIIM).

#### 3.4.2 Model & Training

For training, a Unet model with ResNet50 as the encoder is used. Pretrained weights from ImageNet dataset for encoder is used. For loss function, BCEWithLogits Loss, Dice Loss and Focal Loss were inspected, but Combo loss was used. Adam is selected as the optimizer. During training, augmentations were applied such as scaling and contrasting. 5 fold cross validation was completed. 0.8 to 0.2 ratio was used for training and validation. Several augmentation techniques were applied during training, such as: flipping, random contrast, gamma or brightness and scaling.

## 4. Results

### 4.1 Synthetic Hemothorax Data Framework

This framework processes selected DICOM series and integrates the synthetic objects based on user-defined parameters. Volume calculations are performed to ensure the accuracy and consistency of the generated objects.

The Monte Carlo method for volume estimation was tested on various geometric shapes and under different conditions to assess its accuracy and consistency. The results demonstrate high precision across different object types and sampling rates.

#### 4.1.1 Volume Estimation Accuracy

Table 1 presents the comparison between actual volumes and those estimated using the Monte Carlo method with 1 million samples for different geometric shapes.

**Table 1:**
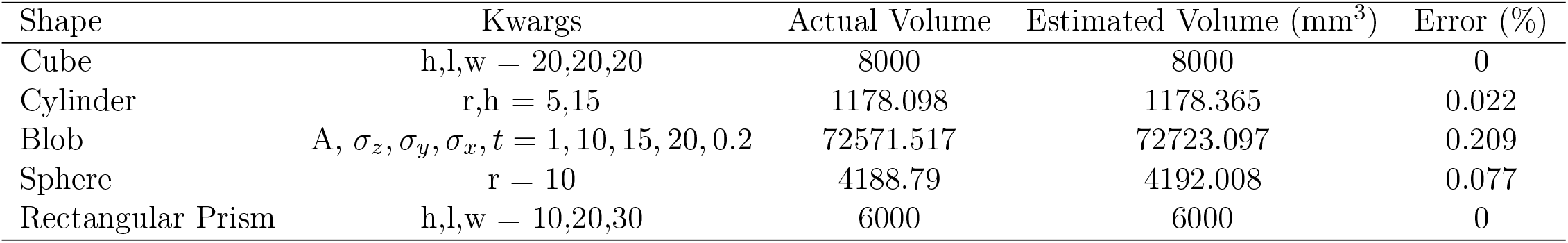
Accuracy of Monte Carlo Volume Estimation.

| Shape | Kwargs | Actual Volume | Estimated Volume ($\text{mm}^3$) | Error (%) |
| --- | --- | --- | --- | --- |
| Cube | h,l,w = 20,20,20 | 8000 | 8000 | 0 |
| Cylinder | r,h = 5,15 | 1178.098 | 1178.365 | 0.022 |
| Blob | A, $\sigma_z, \sigma_y, \sigma_x, t = 1, 10, 15, 20, 0.2$ | 72571.517 | 72723.097 | 0.209 |
| Sphere | r = 10 | 4188.79 | 4192.008 | 0.077 |
| Rectangular Prism | h,l,w = 10,20,30 | 6000 | 6000 | 0 |

The results show excellent agreement between the actual and estimated volumes, with errors consistently below 0.20%. This demonstrates the high accuracy of the Monte Carlo method across different geometric shapes.

#### 4.1.2 Convergence Analysis

To assess the convergence of the Monte Carlo method, we performed volume estimations with increasing numbers of samples for an blob shape, with a base volume of **25.132741*mm***^**3**^’s. Table 2 shows the results.

**Table 2:** Convergence of Monte Carlo Volume Estimation.

| Samples (n) | Volume ($\text{mm}^3$) | Error (%) |
| --- | --- | --- |
| 10,000 | 24.8256 | 1.2220 |
| 100,000 | 25.1356 | 0.0111 |
| 1,000,000 | 25.1172 | 0.0618 |
| 10,000,000 | 25.133357 | 0.0024 |

The estimated volume converges to the actual volume (25.132741) as the number of samples increases, with the estimate at 10 million samples differing by only 0.0024% from the actual value.

#### 4.1.3 Rotational and Scaling Invariance

We tested the method’s invariance to rotation and scaling:

- **Rotation:** The volume of a rotated object differed from the original volume by only 0.19% (25.181376 ***mm***^**3**^, 25.1328 ***mm***^**3**^).
- **Scaling:** When the object was scaled by a factor of 2, the estimated volume (200.552832 ***mm***^**3**^) was within 0.25% of the expected volume (201.062400 ***mm***^**3**^).

These results demonstrate that the Monte Carlo method maintains its accuracy under rotational and scaling transformations, which is crucial for real-world applications where objects may be oriented or sized differently across images.

In summary, the Monte Carlo method for volume estimation shows high accuracy, consistent convergence, and invariance to geometric transformations for various objects, making it a robust tool for volumetric analysis in medical imaging applications.

#### 4.1.4 Volume Estimation

Our results show that volume estimation was validated through test functions, and confirm that the Monte Carlo method used provides accurate and consistent results.

#### 4.1.5 Image Comparison

To illustrate the effectiveness of our framework in integrating synthetic objects into CT images, we present a comparison between original and modified images. Figure 1 shows side-by-side comparisons of original CT scans and the same scans with synthetic objects added.

**Figure 1:**
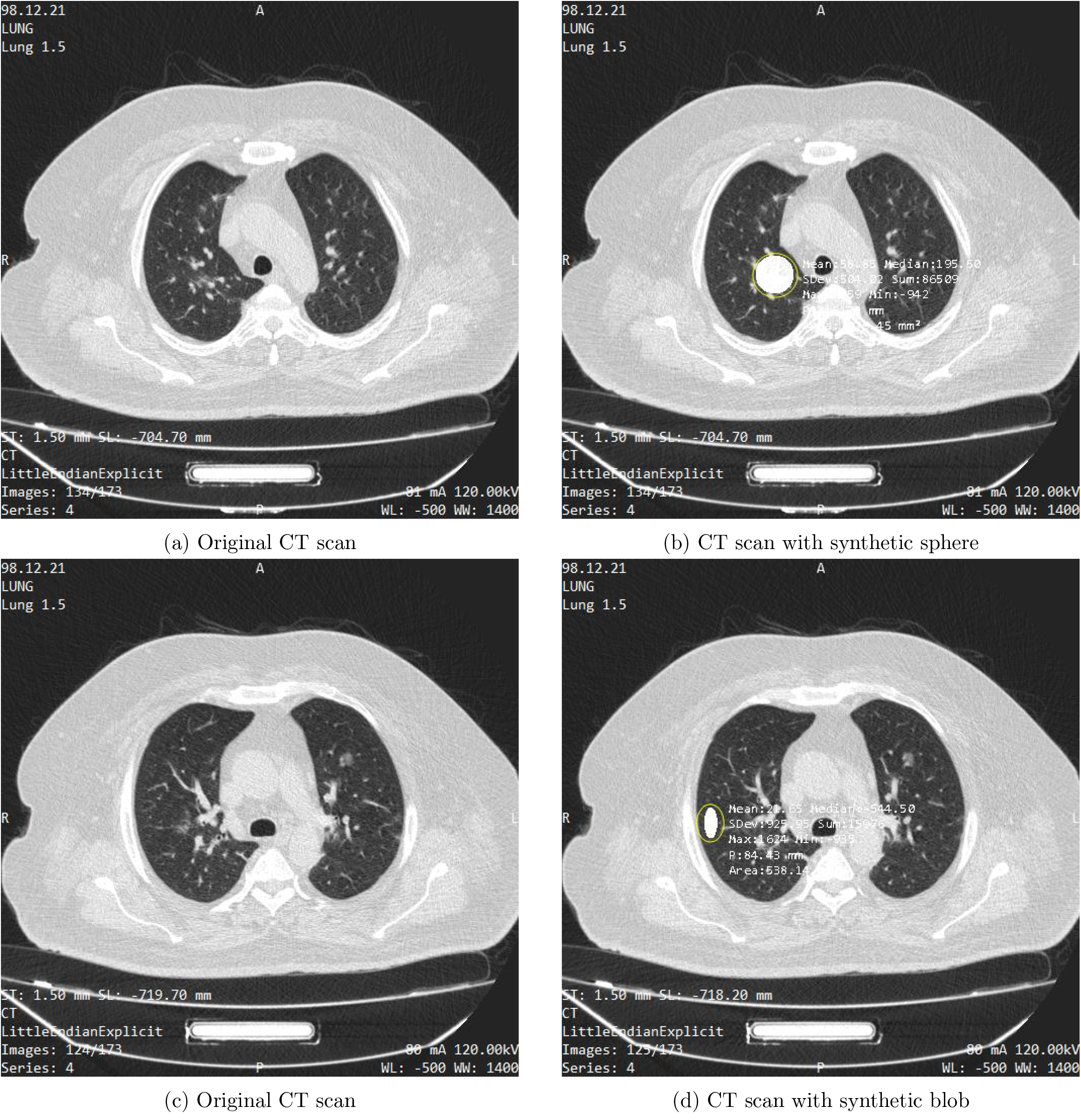
Comparison of original CT scans and scans with integrated synthetic objects.

These images demonstrate the seamless integration of synthetic objects into the CT scans. The added objects (a sphere in Figure 1b and an irregular blob (ellipsoid) in Figure 1d) are clearly visible and realistically incorporated into the thoracic cavity, showcasing the framework’s capability to augment medical imaging data for various analytical and clinical applications with volumetric analysis incorporated.

### 4.2 Deep Learning

#### 4.2.1 Lung, Heart and Trachea

The training was done on a Nvidia (Santa Clara, California, U.S.) P100 GPU 16GB, on Kaggle (San Francisco, California, U.S.) platform. Model was trained for 70 epochs. A 0.8 to 0.2 ratio was used for train and validation datasets.

##### Training Results

Train loss was 0.0152, and validation loss was 0.0233. Train dice score was 0.9622, and validation dice was 0.9537. Train and validation Jaccardi Index scores were 0.9391 and 0.9322 respectively (Jaccard, 1901). F1 scores for lung, trachea and heart are 0.9165, 0.7657 and 0.6458 respectively. The results can be viewed at 2. Example outputs can be viewed at project page. Confusion matrix can be viewed at 4a.

#### 4.2.2 Hemothorax Segmentation & Volume Calculation

The training was conducted on an NVIDIA P100 GPU (16GB), manufactured by NVIDIA Corporation (Santa Clara, California, USA) using the Kaggle platform, which is operated by Google LLC (Mountain View, California, USA). Model was trained for 20 epochs. 0.9 to 0.1 ratio was used for positive and negative object images. 0.8 to 0.2 ratio was used for train and validation datasets.

##### Segmentation Training Results

Results can be seen on the table, average of the 5 runs for each fold. Validation IoU was 0.84 at the end, and loss was 0.0220. No signs of overfitting was observed. The results can be viewed at 3. Example outputs can be viewed at project page. Confusion matrix can be viewed at 4b. F1 score of 0.98 was achieved.

##### Volume Calculation

Linear regression was done on 30 data points, and as hypothesized an R score or 0.999 and ***r***^**2**^ of 0.998 was achieved.

#### 4.2.3 Pneumothorax

The training was done on a Nvidia P100 GPU 16GB, on Kaggle platform. Model was trained for 20 epochs. 0.8 to 0.2 ratio was used for train and validation datasets.

##### Training Results

In average of 5 folds, a Val Dice score of 0.294, loss of 0.77 were achieved. Example outputs can be viewed at project page. The F1 score at the end of the training is 0.36. Confusion matrix can be viewed at 4c.

## 5. Discussion

This study showed that synthetic objects could be used to develop and internally validation a model for assessment of hemothorax, as well as other intra-thoracic lung pathologies. This method could be used for development of other volumetric assessment tools with applications in health care.

**Figure 2:**
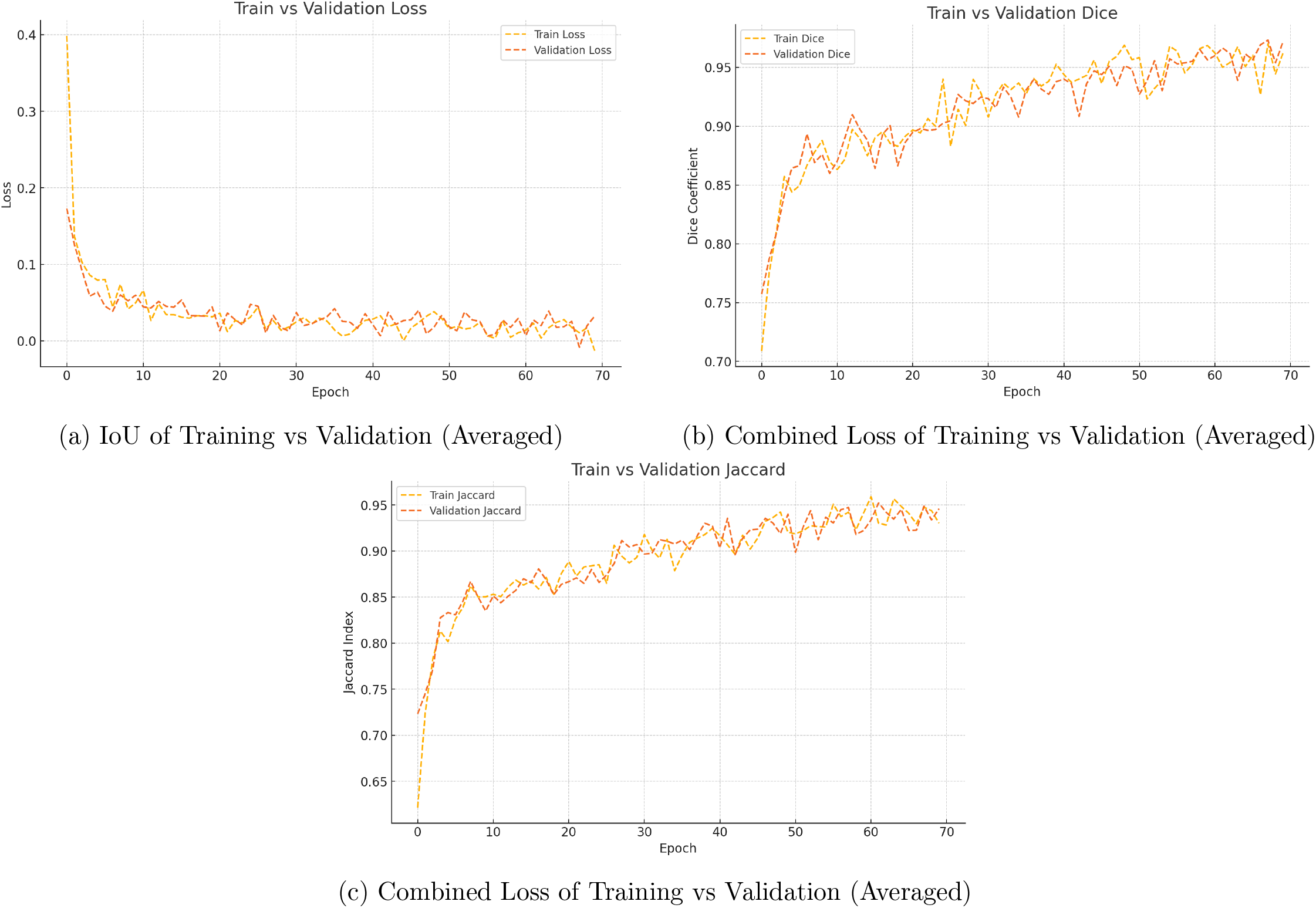
Results of chest model.

**Figure 3:**
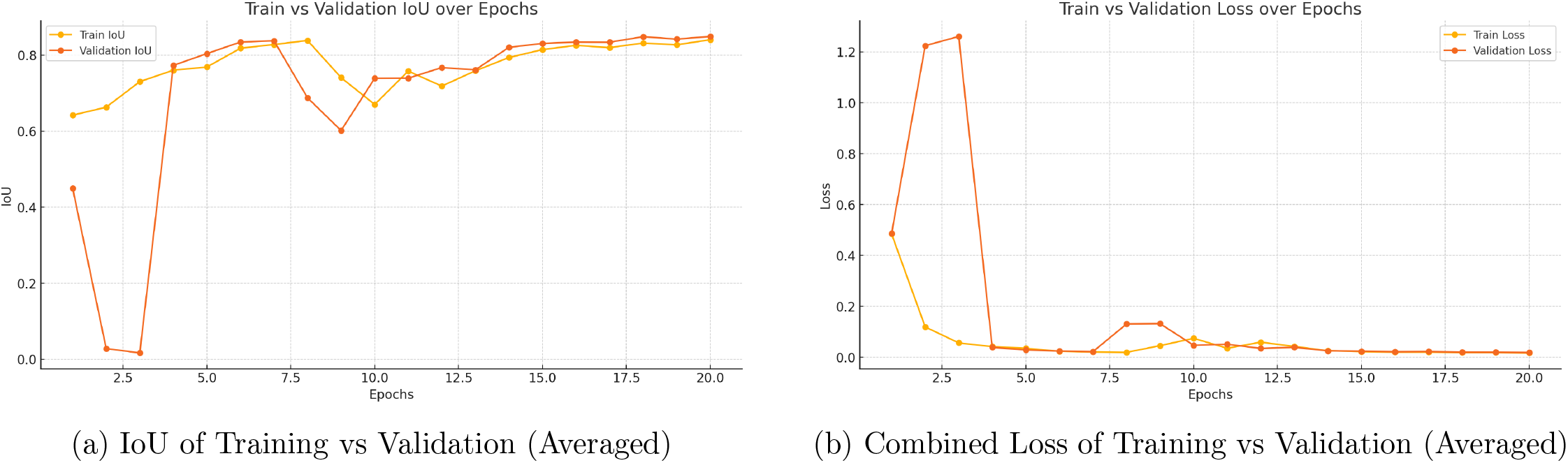
Results of hemothorax model.

**Figure 4:**
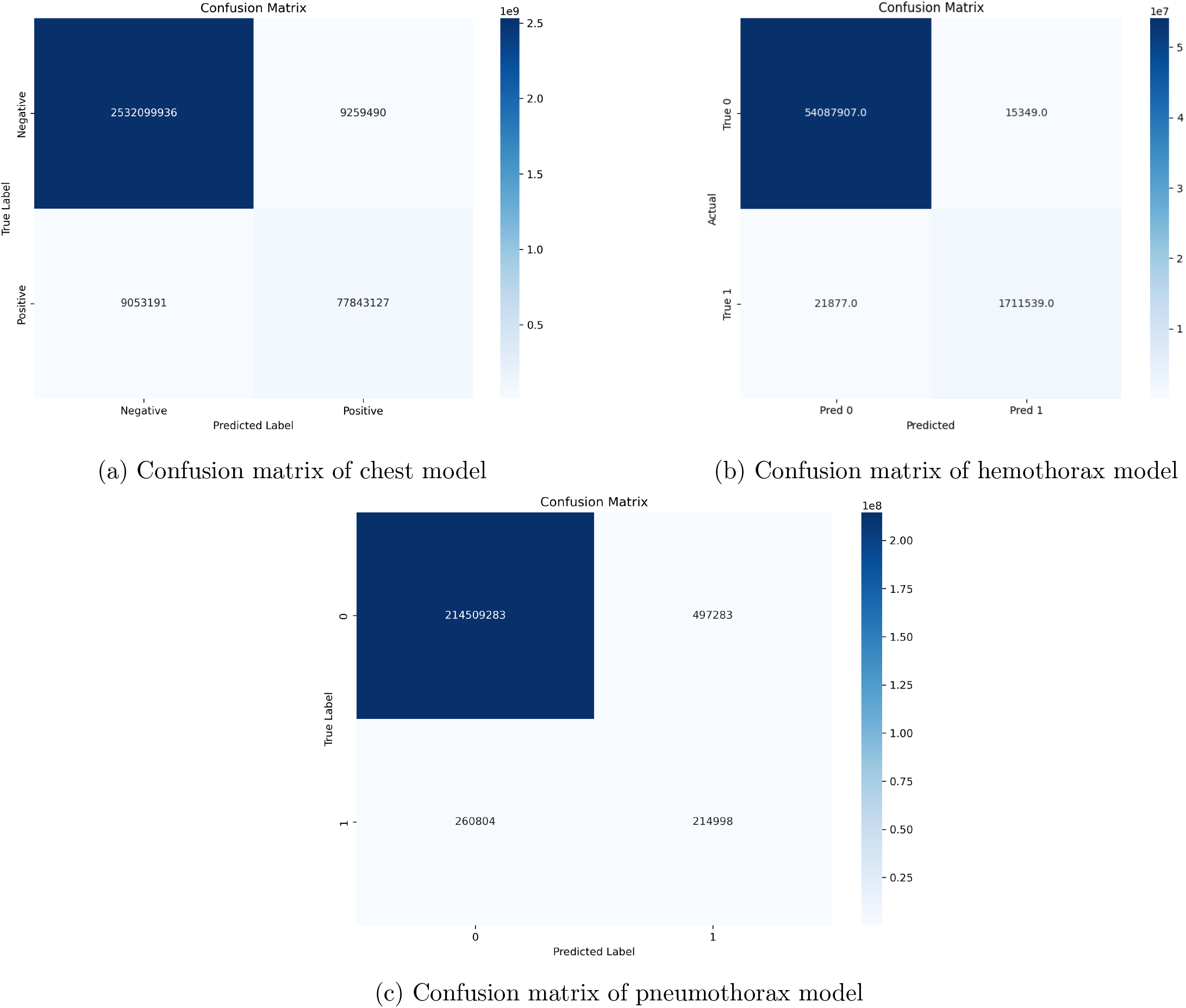
Confusion matrixes.

The integration of synthetic objects into CT imaging data enhances clinical decision making, and can provide a robust tool for data augmentation and analysis, with volumetric assessment. This framework may be able to support various applications in medical imaging and related fields, helping clinicians and surgeons make key clinical decisions.

CT volumetry has been used for the assessment of a variety of pathophysiologies including in the intraabdominal cavity (Planz et al., 2019) as well as the thorax (Nair et al., 2023). However, it has not to our knowledge been used in comparison to determine greater clinical implication compared to other radiographic measures in thoracic trauma imaging.

The framework’s potential is demonstrated using synthetic data created for a CT hemothorax segmentation model. Also, the addition of classification of pneumothorax and the classification of other organ within the mediastinum point to the role of combined use of tools together to create a robust solution for medical analysis.

Next steps include validation of this framework for non-synthetic objects for volumetric assessment in the thorax, and testing hemothorax model with actual data or other synthetic data solutions such as GenerateCT (Hamamci et al., 2024). Further validation will be done externally, testing the model on hospital patients receiving video-assisted thoracoscopy or tube thoracostomy.

Limitations of the study include the lack of external validation of this model, as well as the lack of confirmation of a gold standard for intra-thoracic blood volume. Additional validation could be completed based on comparison of model with the amount of blood volume returned post-procedurally.

## 6. Conclusion

This study presents an initial framework for the integration and analysis of synthetic objects in medical imaging data. The methods and results demonstrate the potential for significant advancements in research and clinical applications.

Additionally, results of UNet models for various pathologies inside thoracic cavity are presented. A potential use case of the framework was exhibited. A possible approach for measuring volume of an object from CT imaging is shown.

## Author Contributions

**First Author**: Conceptualization, Data curation, Methodology, Software. **Second Author**: Conceptualization, Methodology, Writing—original draft.

## Data Availability

Data supporting the findings of this study are available from the corresponding author upon reasonable request. The code for this analysis is available at: [GitHub Repository](https://github.com/ally4n/synthetic_images).

## Acknowledgments

The authors thank the institutions and individuals who contributed to the preparation of this manuscript, as well as the feedback from Dr. Brian Kim and Dr. Daniel Stephens at the Mayo Clinic, Department of Surgery.

